# ClinDataBench: Benchmarking Large Language Model Agents in Clinical Trial Analysis Dataset Construction

**DOI:** 10.64898/2026.11.17.26368316

**Authors:** Zeyu Li, Yichen Luo, Yebo Feng, Jiahua Xu

**Author notes:** **CCS Concepts** - **Computing methodologies** → **Artificial intelligence**; - **Applied computing** → *Health informatics*; - **Software and its engineering** → Software verification and validation.

## Abstract

A new drug reaches patients only after the U.S. Food and Drug Administration (FDA) has reviewed its trial evidence, which every application carries as analysis datasets in the Clinical Data Interchange Standards Consortium (CDISC) Analysis Data Model (ADaM) format that programmers derive by hand from raw trial data, a statistical analysis plan (SAP) and dataset specifications. Coding agents built on large language models (LLMs) could take over this work, and a slow-changing industry will adopt them only with evidence of what they get right and wrong. However, work on large language models (LLMs) for CDISC data stops short of evaluating a complete, conformant and reproducible package, and the standard validator checks structure rather than whether each value is derived correctly. We introduce ClinDataBench, built on the public CDISC pilot study: an agent receives 22 Study Data Tabulation Model (SDTM) domains, the statistical analysis plan (SAP) and specifications, and must deliver 13 Analysis Data Model (ADaM) datasets, Define-XML metadata and a program that rebuilds them offline. Agents run in containers whose only network route is their model API, restricting online access to published reference outputs during execution. A host-only evaluator scores conformance under the Pinnacle 21 Community (P21) validator, selected derivations, clean reconstruction and effort separately. We compare three settings: Base, which submits the agent’s first package; Self-Refine, which revises it without external validation feedback; and CRITIC, which revises it with the agent’s own validator report. In an exploratory pilot with six models, the three closed models and DeepSeek delivered all 13 datasets in every setting and eleven of their twelve packages rebuilt semantically, while two other open-weight models delivered no complete package. Validator findings ranged from 15 to 64 dataset-rule pairs. Four of six CRITIC workflows had usable validator comparisons, and each reduced its finding count. None reduced its failed derivation count. The package with the fewest findings carried none of the 234 efficacy summary records in the form the specifications define, showing that lower validator finding counts do not necessarily indicate more correct derivations.

## 1 Introduction

A new drug reaches patients only after regulators have reviewed its trial evidence, and the wait has a measurable price. Faster review by the U.S. Food and Drug Administration (FDA) under the Prescription Drug User Fee Acts saved an estimated 140,000 to 310,000 life-years [22], and a single day of delay in drug development is worth about $800,000 in lost prescription sales, on top of about $40,000 in direct daily trial costs [28]. Reviewers reanalyse the trial data themselves, so every submission must ship its data in a standard form they can recompute from. The FDA requires the standards of the Clinical Data Interchange Standards Consortium (CDISC) [31]: standardized records in the Study Data Tabulation Model (SDTM) and derived, analysis-ready datasets in the Analysis Data Model (ADaM) [4].

Programmers build the ADaM datasets by hand. They read the trial’s statistical analysis plan, write the code that derives each analysis variable, such as a baseline value, a visit-window average or the time to an adverse event, and check the output with a rule-based validator before submission [33]. Most of the industry verifies each dataset by double programming, in which a second programmer derives it again independently [9]. The datasets can also sit directly on the path to review: under the FDA’s Real-Time Oncology Review, sponsors send key raw and ADaM datasets ahead of the full application, a median of 5.7 weeks earlier, so that review starts sooner [8, 32]. Figure 1 places the task in the submission pipeline, and Table 1 maps each clinical term to its data-engineering counterpart.

**Table 1:**
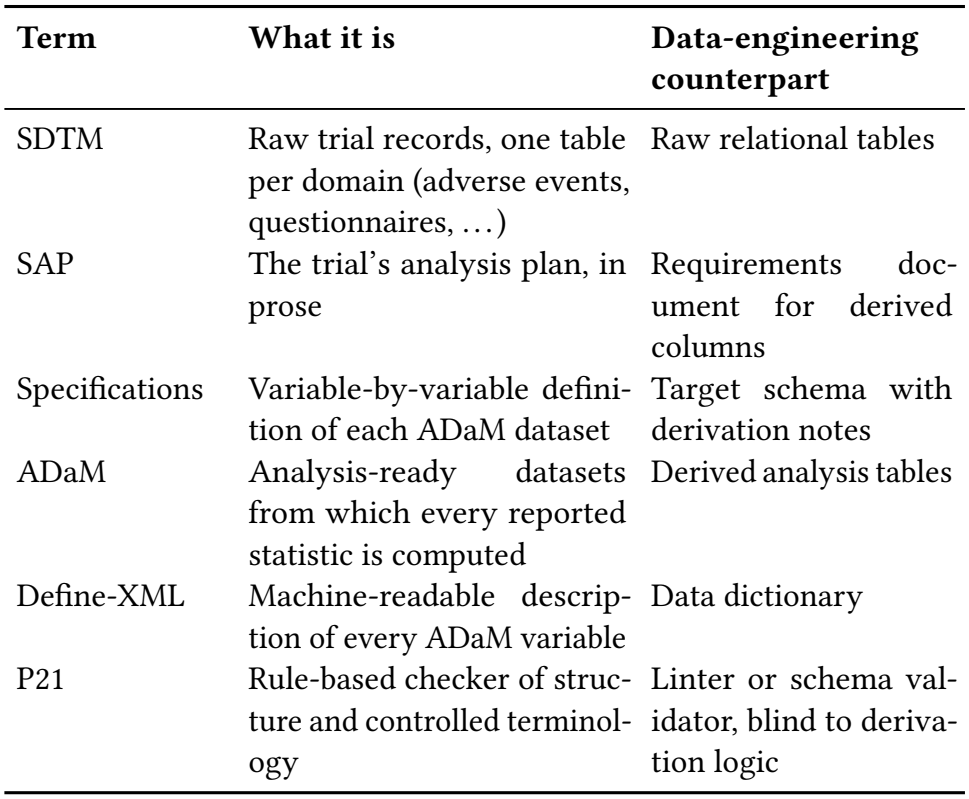
ClinDataBench terms and their closest data-engineering counterparts.

| **Term** | **What it is** | **Data-engineering counterpart** |
|---|---|---|
| SDTM | Raw trial records, one table per domain (adverse events, questionnaires, …) | Raw relational tables |
| SAP | The trial’s analysis plan, in prose | Requirements doc-ument for derived columns |
| Specifications | Variable-by-variable defini-tion of each ADaM dataset | Target schema with derivation notes |
| ADaM | Analysis-ready datasets from which every reported statistic is computed | Derived analysis tables |
| Define-XML | Machine-readable descrip-tion of every ADaM variable | Data dictionary |
| P21 | Rule-based checker of struc-ture and controlled terminol-ogy | Linter or schema val-idator, blind to deriva-tion logic |

**Figure 1:**
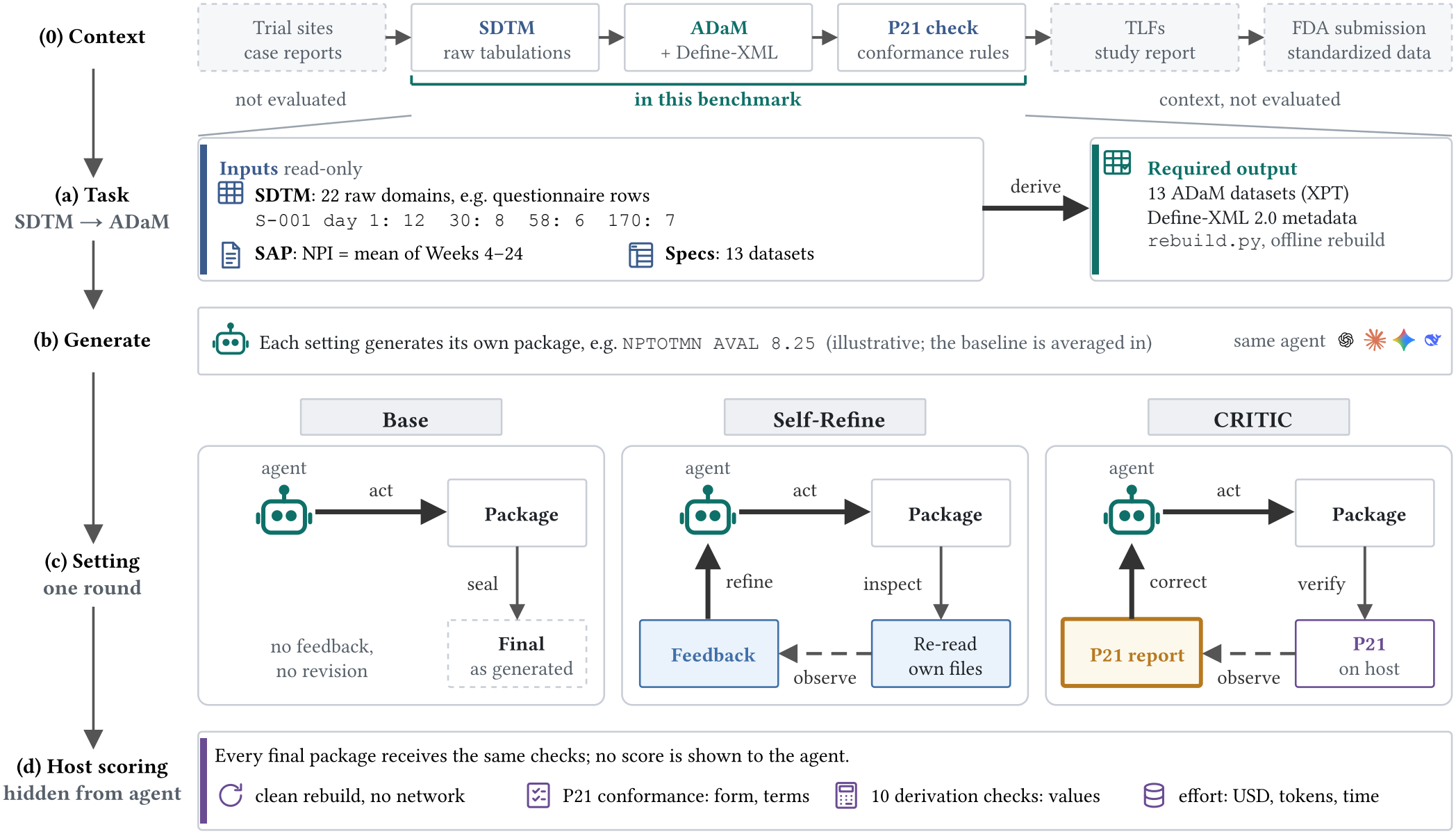
ClinDataBench overview. (0) Context: in a drug submission, tabulated trial data (SDTM) become the analysis datasets (ADaM) from which the study report is computed, and the FDA asks sponsors to check study data against CDISC and FDA rules before submission [33]; the benchmark covers the bracketed steps only. (a) Task: from read-only SDTM domains, the SAP and the specifications, the agent must deliver 13 ADaM datasets, Define-XML and an offline rebuild program. (b) Each setting generates its own package with the same agent and prompt; the row shown is illustrative. (c) Settings: Base submits the generated package; Self-Refine revises it after feedback on its own files; CRITIC revises it after its own Pinnacle 21 Community (P21) report (highlighted), which the host runs once. (d) A host-only evaluator scores every final package on the same checks and shows no score to the agent.

We ask whether coding agents built on large language models (LLMs) can deliver complete, reproducible ADaM data packages that satisfy explicit specifications. We further ask whether validator-guided revision improves analytical correctness alongside conformance. The bar for adoption is high, because the industry changes its tools slowly. FDA has not required any specific statistical software since 2015 [30], yet the first public test submission whose ADaM datasets were built in R received its final FDA response only in August 2024 [23], and every dataset is still submitted as a SAS XPORT version 5 file [33]. An agent will enter this workflow only with evidence of what it gets right and what it gets wrong.

Coding agents now resolve real software issues [15] and write data-science code [16]. A parallel line of work studies how such models correct themselves: LLMs often fail to improve when asked to review their own answers [13], while feedback from external tools does help [7, 11]. ADaM programming looks like a natural next target for both lines. Its inputs are tabular, its rules are written down, and an automatic validator already exists to supply external feedback.

However, work on LLMs for CDISC data stops short of evaluating a regulatory package. An agent architecture reports the structural match of two ADaM datasets from the same pilot study [37], industry pipelines report proof-of-concept results without a cross-model evaluation [1, 17], and agent toolkits leave Define-XML and Pinnacle 21 Community (P21) compliance to future work [36]. These studies leave three evaluation requirements unresolved. The validator checks structure and controlled terminology, not whether a baseline was chosen from the right visit; on oncology submissions, its FDA engine detected 6 of 20 contradiction archetypes [35], so a low finding count can reward a wrong package. The reference outputs of the public CDISC pilot study are published online, and search agents already retrieve benchmark answers from the web [12], so an agent with web access could copy them. And a package is useful only if it can be rebuilt from its programs, which a single score of the output files does not test. The validator also leaves the self-correction question open: its report is external feedback, but it speaks only to conformance, and whether it helps an agent improve a package or merely lowers its finding count has not been measured.

To fill this gap, we introduce ClinDataBench, a benchmark in which an agent transforms a real trial’s SDTM domains, statistical analysis plan (SAP) and dataset specifications into an ADaM data package comprising analysis datasets, Define-XML metadata and executable reconstruction programs. The benchmark evaluates conformance, selected derivations and reproducibility separately, while restricting online access to published reference outputs during execution. It also lets us compare, on the same task, a revision without external validation feedback, Self-Refine [19], with a revision guided by the agent’s own validator report, CRITIC [11].

ClinDataBench has four parts (Figure 1). The task package, built from the CDISC pilot study, gives the agent 22 SDTM domains, the SAP and the specification workbook, and asks for 13 ADaM datasets, Define-XML metadata and a program that rebuilds them offline (section 3). The sandbox runs each agent in a container whose only network route is an allowlisted gateway to its model API. The settings protocol has every workflow generate once under a shared prompt and seals the package before the agent learns whether it stops there (Base), revises without external validation feedback (Self-Refine) or revises with its own P21 report (CRITIC) (section 4). The host-only evaluator scores conformance under the P21 validator, ten selected derivation checks recomputed from the SDTM inputs, clean reconstruction and effort, and never combines them into one number.

In an exploratory pilot, we evaluate three closed models, `gpt-6.1-sol, claude-sonnet-5-5` and `gemini-3.1-pro-preview,` and three open-weight models, `deepseek-flash, gpt-oss-20b` and `llama-4-scout,` in each of the three settings (section 5). The closed models and `deepseek-flash` delivered all 13 required datasets in every setting, and eleven of their twelve packages rebuilt semantically in a clean container; the other two open-weight models delivered no complete package. The same six models also ran in each architecture extension. In the pilot, validator findings ranged from 15 to 64 dataset-rule pairs. All four CRITIC workflows with usable reports reduced their finding counts. None reduced its failed derivation count. The package with the fewest findings, 15 pairs, carried none of the 234 Week 4–24 Neuropsychiatric Inventory (NPI) summary records in the form the specifications define, while all three `gpt-6.1-sol` packages and the `deepseek-flash` CRITIC package, with 24 to 26 pairs, passed all ten derivation checks.

Our paper makes three contributions:

- **A benchmark for regulatory analysis datasets**. ClinDataBench pairs a public trial package with an isolated sandbox and a hostonly evaluator, so a package is scored on conformance, selected derivations, reconstruction and effort while restricting online access to published reference outputs during execution.
- **A protocol for evaluating self-correction**. Shared generation prompts and sealed checkpoints keep generation uninformed of the assigned setting. Within-workflow comparisons and a content guard track whether reductions in validator findings coincide with improvements in selected derivations or losses of content.
- **Evidence that lower finding counts need not mean more correct derivations**. In a six-model pilot, the package with the fewest validator findings failed selected derivation checks that packages with more findings passed, so this domain needs derivation checks next to the validator.

## 2 Related Work

### Benchmarks for coding and data agents

SWE-bench scores agents by whether repository tests pass after their patch [15]; DS-1000 checks data-science snippets against executed test cases [16]; MLE-bench grades complete machine-learning pipelines on held-out Kaggle metrics [6]; AgentBench spans operating systems, databases and web environments [18], and Terminal-Bench runs agents on tasks inside containers [20]. PaperBench additionally requires a submission to reproduce its results from scratch in a fresh environment [29]. Each defines success by one kind of automatic signal. ClinDataBench instead has two signals that can disagree, a regulatory validator and recomputed derivations, and it adds clean reconstruction as a third requirement. We run all agents through one open agent harness, OpenCode [21], in the spirit of open platforms such as OpenHands [34], so that model differences are not confounded with differences between vendor applications.

### Self-correction and external feedback

ReAct interleaves reasoning with tool observations [38], while Reflexion [26] and Self-Refine [19] let a model critique and revise its own output. Without an external signal, such revision often fails to improve reasoning [13]; with tool feedback, from a search engine, an interpreter or unit tests, it does [7, 11]. These studies compare feedback sources on short tasks with an unambiguous answer. Our Self-Refine and CRITIC settings differ by one external report on a long task, and the report measures only one of the properties the task requires.

### LLMs in clinical research

LLMs answer medical examination questions [27] and act as agents in virtual electronic health records [14]. TrialAgentBench asks agents to analyse patient-level trial data, choose statistical methods and make drug-development decisions [25]. ClinDataBench addresses an earlier stage of the pipeline: not the analysis or the decision, but the regulated data engineering that turns tabulated trial records into the analysis datasets those analyses consume.

### LLMs for CDISC data

GxP-Agent organises LLM agents as a process graph for clinical trial programming and reports the structural match of ADSL and ADAE generated from the same pilot study [37]. ClinAgent packages agent skills for ADaM and table programming and lists Define-XML generation and P21 compliance as future work [36]. Industry papers describe a multi-agent ADaM pipeline that regenerates code after execution errors [17] and a loop that revises LLM-generated SDTM mappings from CDISC CORE rule violations [1], without a cross-model evaluation. CAVE-Onc shows that the P21 and CORE rule engines miss most contradiction archetypes in oncology submissions [35]. ClinDataBench asks for the complete 13-dataset package with Define-XML and a rebuild program, scores it with the regulatory validator and recomputed derivations side by side, and compares revision with and without the validator report.

### Contamination

Public benchmark items can enter training corpora or be retrieved at test time [10, 24], and search agents have found ground-truth labels online for about 3% of questions on three benchmarks [12]. Our inputs are public, so we cannot rule out that a model saw the pilot study in training. We remove retrieval at test time instead: the agent cannot reach the web.

## 3 The ClinDataBench Task

### 3.1 Study Data

We build the task from the CDISC SDTM/ADaM Pilot Project [2], which converted de-identified legacy data from a phase II trial of xanomeline in mild to moderate Alzheimer’s disease into a CDISC submission. The trial randomized patients to placebo, low dose or high dose. CDISC distributes the study with its SAP, SDTM tabulations and reference ADaM datasets. We give the agent only the first two; the reference ADaM datasets are never supplied.

The agent package has three parts (Figure 1a). The 22 SDTM domains arrive as SAS transport (XPT) files, which are authoritative; the trial summary domain is encoded in Windows-1252, and the prompt says so. The SAP arrives as a PDF. The specification workbook arrives as a JSON export of its cell values, with formulas rejected at preparation. The specifications define 13 ADaM datasets: subject level (ADSL), adverse events (ADAE), concomitant medications (ADCM), disposition (ADDS), exposure (ADEX), three laboratory datasets (ADLBC, ADLBH, ADLBHY), vital signs (ADVS), time-to-event (TTE) (ADTTE), and three questionnaire datasets for ADAS-Cog, CIBIC+ and the NPI (ADQSADAS, ADQS-CIBC, ADQSNPIX).

We found one internal contradiction in the original workbook. It declares the vital-signs treatment variable ADVS.TRTA as character but names the numeric ADSL.TRT01AN as its source. We correct the source to the character ADSL.TRT01A in the exported specification. This is the only change; we record the workbook hash and the cells before and after in a provenance file the agent never sees.

### 3.2 Deliverable Contract

The agent must write, under one output directory, the 13 datasets as lowercase XPT files, a Define-XML 2.0.0 file describing them, and the programs that produce them, with a single entry point `rebuild.py`. Running that program in a clean workspace with the same read-only inputs and no network must regenerate every dataset and the Define-XML. The agent also writes a manifest and notes that document source lineage, assumptions, dependencies and incomplete derivations. The full generation prompt is in Appendix A.

The contract makes three demands that a score of the output files alone would miss. The package must be complete, it must be described by metadata a reviewer can read, and it must be reproducible from code. A regulator needs all three.

### 3.3 Isolated Execution

Each workflow runs in its own Docker container with 4 CPUs, 8 GB of memory, a 256-process limit, no Linux capabilities and a non-root user. The container mounts only three things: the read-only inputs, a writable workspace, and a private volume that holds the agent’s session history. Controller code, scores, reference datasets and the validator are absent.

We restrict the network inside the container’s own namespace. An allowlisting gateway forwards HTTPS only to the assigned provider’s API host, checks the TLS server name against that host, and rejects everything else. IPv4 and IPv6 rules drop all other outbound traffic, including DNS and UDP. Before each workflow, probes confirm that direct HTTP, HTTPS and DNS fail and that a non-provider host is refused; a failed probe stops the workflow before any model call. The agent’s web-fetch, web-search and sub-agent tools are disabled, and package installation is unavailable. The agent therefore cannot retrieve the published reference datasets, CDISC documentation or the validator during a run. The gateway does not decrypt traffic, so it cannot inspect what the agent sends to its own provider; the prompt prohibits provider-side search tools.

## 4 Settings and Evaluation

### 4.1 Three Settings

ClinDataBench compares three settings, each named after the method it follows. *Base*, the reference condition of Self-Refine [19], is the agent’s generation phase alone: the OpenCode agent reads the inputs, writes and runs its programs and inspects the outputs over many tool calls, then submits the package without a revision turn. In *Self-Refine* [19], the agent then gives feedback on its own programs, datasets, metadata and logs against the inputs and refines the package; unlike the original method, it stops after a single round. In *CRITIC* [11], the agent instead revises the package after an external tool verifies it: its own P21 report; unlike the original method, where the model calls the tool itself, the controller runs P21 once and delivers the report. All three settings run the same OpenCode agent loop; Self-Refine and CRITIC add one revision turn and differ only in whether it receives the report. Every revised workflow has two phases in one persistent OpenCode session (Figure 1c).

1. **Generation**. The agent receives the generation prompt. The prompt is identical in all settings, and nothing in the container reveals the setting. There is no time cap.
2. **Checkpoint**. When the agent ends its turn, the controller pauses the container, copies and hashes the workspace, and evaluates the copy on the host.
3. **Revision**. The controller resumes the same session by its identifier and sends the revision prompt of the assigned setting. In the CRITIC setting it first copies the initial report into the workspace. If no usable report exists, that workflow receives a standard report-unavailable message, continues as a Self-Refine revision and stays in the CRITIC setting. The controller verifies that the exported session history preserves and extends the history from generation.
4. **Final checkpoint**. The controller seals and evaluates the revised workspace exactly as in step 2.

The two revision prompts share every sentence but the last (Appendix A). Both tell the agent that P21 assesses conformance and does not establish analytical correctness, and both forbid running any validator. The Self-Refine prompt ends with “No external validation feedback is available during your review”; the CRITIC prompt ends with the location of the report. No derivation-check result reaches either setting.

We keep generation independent across settings. A paired design would fork one initial package into both revision settings, but then the CRITIC setting’s generation would be the same draw as the Self-Refine setting’s, and a fork would have to discard a session the agent already holds. We run instead 18 independent workflows in the main study, three models × two revision settings × three repetitions, in an order shuffled by a fixed seed. Every workflow gets its own generation and exactly one revision. The controller never retries a phase, swaps a model or stops early on a score; a failed workflow halts the schedule until it is adjudicated, and it remains in the denominator.

### 4.2 Evaluation Metrics

A host-only evaluator scores both checkpoints of every workflow on four metric groups (Figure 1d). It reads copies of the sealed packages and the raw SDTM inputs; nothing it computes enters the agent container except, in the CRITIC setting, the initial P21 report. A metric that cannot be computed is recorded as unavailable, never as zero.

#### Delivery and reproduction

**Data** counts the required datasets that exist and can be read, out of 13. To test reproduction, we copy only the agent’s programs into a fresh container without network and run rebuild.py with a limit of 1,800 seconds. **Semantic reproduction** (*Sem*.) checks whether every rebuilt dataset has the same values, row order, labels, types and storage widths as the delivered one, and whether the canonicalized Define-XML is identical up to its creation timestamp. **Byte reproduction** (*Byte*) checks whether every file hash matches.

#### Conformance

The FDA asks sponsors to evaluate their study data against CDISC conformance rules and FDA business rules before submission [33], and P21’s FDA engines carry the validation rules of DataFit, the FDA’s own implementation of Pinnacle 21 [5]. A conformance score in ClinDataBench is therefore the check a sponsor would run before submitting the package. We use the P21 command-line client 1.0.9 with the FDA 2508.1 engine, ADaMIG 1.3 [4], the ADaM 2.1 model [3], CDISC controlled terminology of 2026-03-27 and English MedDRA 8.0, with the DM, AE and EX domains as supporting SDTM data. Every call runs on a disposable copy of a pinned runtime cache whose hashes are frozen.

Let *D* be the delivered datasets, *R* the validator’s rules, and *n*_*d,r*_ the number of findings that rule *r* raises on dataset *d*. The primary conformance metric is the number of **dataset-rule pairs** with findings,

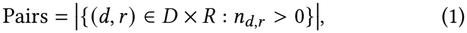

which counts a rule once per dataset however often it fires. We also report **finding stances** (*Inst*.), Σ_*d,r*_ *n*_*d,r*_, and **distinct rules** (*Rules*), |{*r* : *Σ*_*d*_ *n*_*d,r*_ *>* 0}|. We require complete deliverables and validation coverage of all 13 required datasets for the primary conformance score. CRITIC can use a report for feedback if it is fresh and parseable, reports the pinned versions, shows no installation problem, and validates a nonempty subset of the produced datasets. Counts from partial reports describe only the datasets validated.

A drop in pairs can come from deleting the records that triggered the findings. A content guard therefore compares the two checkpoints of each workflow and flags any removed column, lost row, fewer nonmissing values, lost subject or subject–parameter combination, and any new error or smaller denominator in the NPI checks. We count a reduction in pairs as a *guarded improvement* only if the guard raises no flag and all its evidence is available.

#### Selected derivations

The validator does not recompute derivations, so we recompute ten of them from the raw SDTM data on the host and compare them with the delivered datasets, with relative tolerance 10^−10^ and absolute tolerance 10^−8^. The checker reads only XPT files and never runs the agent’s programs. Three checks cover the secondary efficacy endpoint, the mean NPI total score over Weeks 4–24: whether every randomized subject has exactly one summary record, whether its value equals the mean of the scores selected inside the SAP visit windows, and whether its baseline equals the score closest to Day 1. Seven checks cover the time to the first dermatologic adverse event in ADTTE: subject coverage of the safety population, start date, event status, the link from each event to its ADAE record, event date, duration arithmetic, and the censoring date. Event checks are conditional on the treatment-emergent and dermatologic flags in the agent’s own ADAE, and the duration and censoring checks test conventions the SAP does not spell out; Appendix B lists every rule.

From these checks we report three metrics. **Scored** counts the checks that could be scored, out of 10; a check is unscorable when the records it needs are absent. **Failed** counts the scorable checks with at least one wrong record. **Coverage defects** (*Cov*.) sum the missing, extra and duplicate subjects in the two coverage checks.

#### Effort

From the agent’s event stream we record the cost Open-Code estimates for each step (*USD*), provider-reported input and output tokens, including cache reads and writes, agent minutes, and tool calls.

## 5 Pilot and Architecture Results

### 5.1 Pilot Design

Before the main study, we ran an exploratory pilot that is excluded from it, with three closed models, `gpt-6.1-sol, claude-sonnet-5-5` and `gemini-3.1-pro-preview`, at provider-default sampling settings, and three open-weight models. deepseek-flash is DeepSeek’s API alias, which served V4.1-Flash-1017 at configuration time; DeepSeek releases these weights under the MIT license, although the alias does not guarantee that the served weights match the release.`gpt-oss-20b and llama-4-scout` ran through a hosted inference provider at temperature 1.0, with medium reasoning effort for `gpt-oss-20b;` hosted inference is not a private local deployment. Each model ran in a separately frozen extension with the same inputs, prompts, image and evaluator. We ran each model once per setting. Table 2 reports all 18 pilot workflows and all 12 architecture attempts. An audit confirmed for each workflow that checkpoints were unaltered, the session continued unbroken, the requested model answered, and the prompts matched the frozen versions.

**Table 2:**
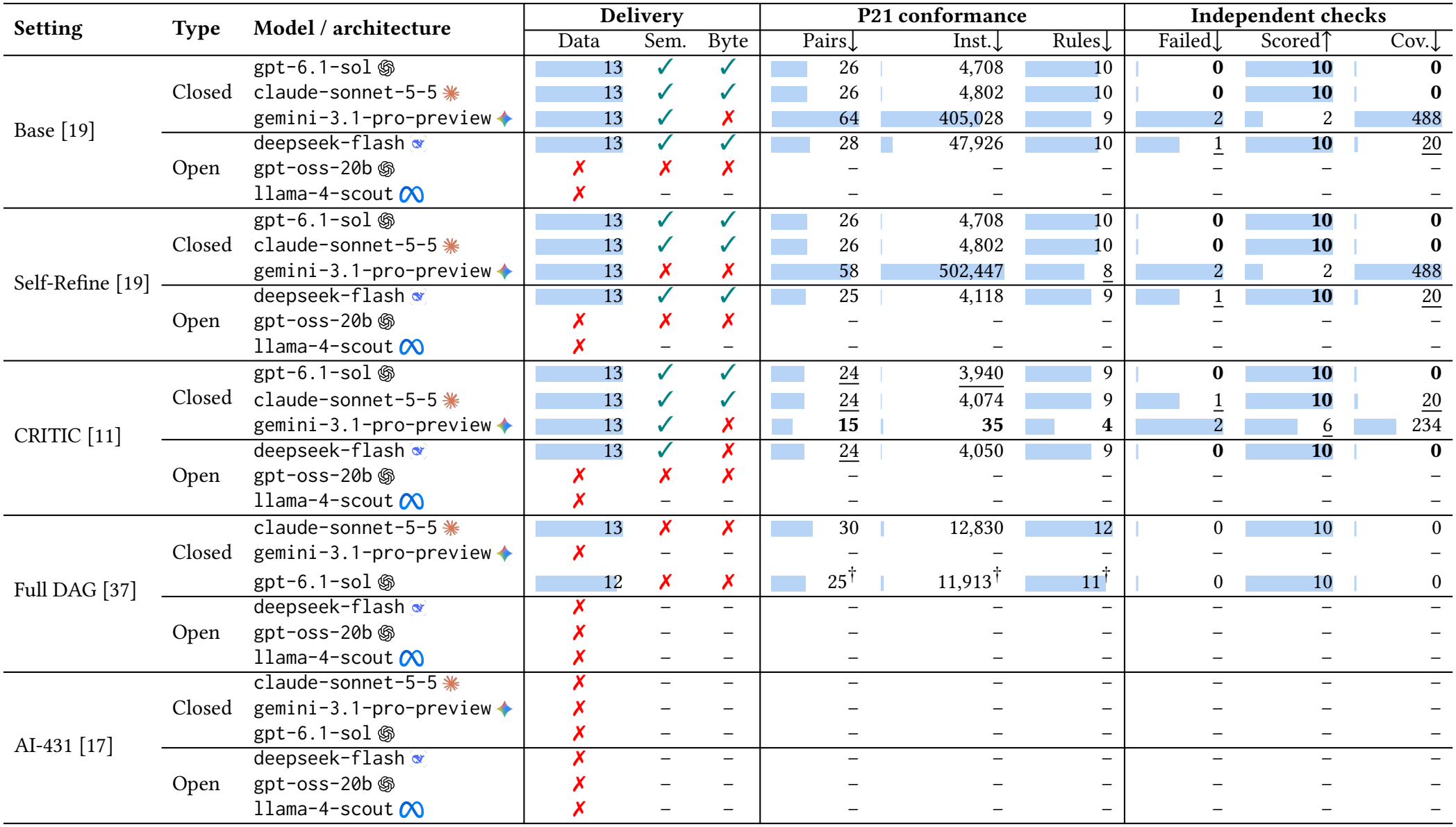
Excluded exploratory results; subsection 4.2 defines every metric. The upper panel reports the 18 pilot packages: *Base* precedes revision, while *Self-Refine* and *CRITIC* follow one revision without and with the agent’s own P21 report. The lower panels report the full-benchmark GxP-Agent-inspired process DAG and AI-431-inspired variable pipeline. Both architectures target 13 datasets and use the ten checks. The historical Claude DAG row includes a storage-only continuation; the fresh gpt-6.1-sol DAG row delivered a partial package, so its daggered P21 counts are descriptive. A cross under *Data* marks no accepted package; a dash is unavailable. Architecture attempts are not ranked. Over the 18 pilot rows, bold marks the best and <u>underline</u> the second-best value, ties included. Every numeric cell has a bar scaled to the full-table maximum of its column; a thin line marks zero. The content guard excludes three content-losing revisions from improvement claims (subsection 4.2).

### 5.2 Delivery and Reproduction

The packages of the three closed models and `deepseek-flash con-` tained the 13 required datasets in readable form, together with the Define-XML, manifest, notes and rebuild program. The other two open-weight models delivered no complete package in any setting: their sessions ended normally after seconds to about a minute of agent time, without the required datasets, so no P21 report or derivation check could be scored. Their CRITIC workflows therefore received no usable report and fell back to the Self-Refine prompt. The failures were early stops rather than wrong derivations: `llama-4-scout` spent 7 to 24 seconds of agent time per workflow, and the validator found no Define-XML or inputs to check. The `gptoss-20b` Base run wrote only rebuild scaffolding; its Self-Refine run looked for the inputs under a relative inputs path instead of the mounted `/inputs,` and its revision turn made no tool call at all. File delivery alone hid schema omissions. Ten of the twelve complete packages matched all 531 required dataset–variable name/type pairs; the `gemini-3.1-pro-preview` Base and Self-Refine packages matched 422 and 436, despite delivering all 13 readable files. The six incomplete open-weight packages delivered 0 of 13 datasets and matched 0 of 531 pairs (Appendix C). Of the twelve complete packages, eleven reproduced semantically in a clean container and eight byte for byte; none of the three `gemini-3.1-pro-preview packages` and not the`deepseek-flash` CRITIC package reproduced byte for byte. The `gemini-3.1-pro-preview` Self-Refine package did not reproduce at all: its rebuild program exited normally but regenerated ADQSNPIX with 71,200 rows instead of the 61,450 it had submitted, while the other 12 datasets matched.

### 5.3 Architecture Extensions

Both architectures target 13 datasets and 531 variables with a 32,768-token output cap. The process DAG has 73 nodes, two attempts per node and two dataset branches. Its historical `claude-sonnet-5-5` continuation delivered all 13 datasets and passed all ten selected derivation checks, but did not reproduce semantically. A fresh `gpt-6.1-sol` attempt passed 72 nodes, delivered 12 datasets with 494 variables and passed all ten checks. Its partial P21 report contained 25 dataset–rule pairs, and neither reconstruction test passed. The fresh `gpt-oss-20b` and `llama-4-scout` attempts were rejected on their first clinical requests because the requested contexts exceeded the provider’s 131,072-token limit; earlier `gemini-3.1-pro-preview` and `deepseek-flash` attempts stopped before export.

The AI-431-inspired pipeline generates each variable independently, executes and repairs it locally, then applies keyed merging and cross-variable QC. None of six clinical attempts produced an accepted dataset. `claude-sonnet-5-5` and `deepseek-flash` each passed all 49 ADSL variable tasks before inconsistent treatment durations blocked the merge. The other runs passed 30 tasks with `gemini-3.1-pro-preview,` 37 with `gpt-oss-20b` and 48 with `gpt-6.1-sol,` whose last request timed out. `llama-4-scout` passed none: 48 tasks exhausted both attempts and VISNUMEN ended with a 300-second transport timeout. The architecture results are exploratory attempts with different providers and execution histories, not model rankings.

### 5.4 Conformance

All twelve complete pilot packages had a usable P21 report. The Base packages carried 26, 26, 64 and 28 dataset-rule pairs. Self-Refine left the `gpt-6.1-sol` and `claude-sonnet-5-5` packages unchanged in pairs and finding instances, at 26 pairs with 4,708 and 4,802 instances. For `gemini-3.1-pro-preview`, it removed 16 pairs and introduced 10, a net change from 64 to 58, while finding instances rose from 405,028 to 502,447. For `deepseek-flash`, it removed 3 pairs, from 28 to 25, and cut finding instances from 47,926 to 4,118. The four CRITIC workflows with usable reports removed 2, 2, 34 and 23 pairs and introduced none, ending at 24, 24, 15 and 24.

Figure 2 shows the eight OpenCode pilot revisions with paired reports. Among the open-weight models, `deepseek-flash` resolved 3 pairs in Self-Refine and 23 in CRITIC. Neither `gpt-oss-20b nor llama-4-scout` delivered the files needed for paired reports. `gemini-3.1-pro-preview` Self-Refine was the only revision to introduce new pairs. The architecture extensions report terminal P21 outcomes in Table 2.

**Figure 2:**
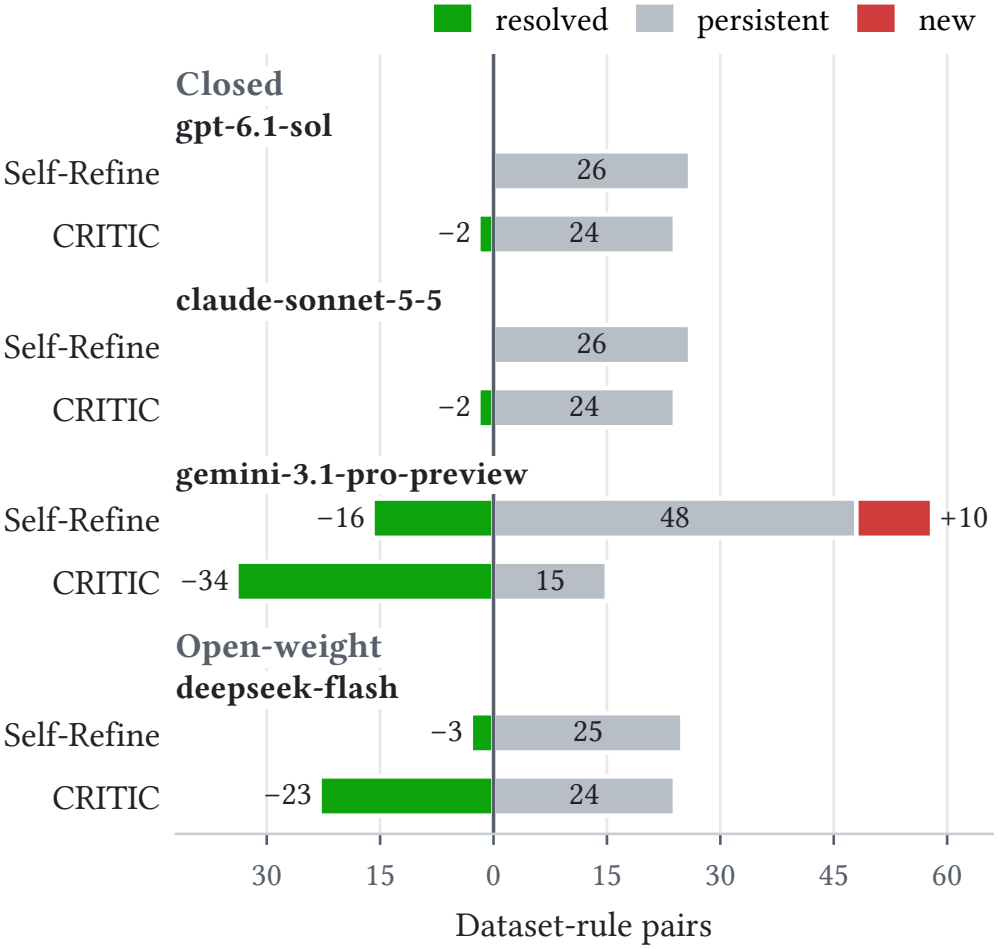
P21 transitions for OpenCode pilot revisions with paired reports.

Three reductions are guarded improvements: the `claude-sonnet-5-5` CRITIC revision and both `deepseek-flash` revisions. The content guard raised 6 and 77 flags on the `gpt-6.1-sol and gemini-3.1-pro-preview` CRITIC revisions and 20 on the `gemini-3.1-pro-preview` Self-Refine revision, so those reductions may partly reflect lost content rather than corrected content.

Two changes to the validator procedure preceded these scores. Earlier runs had logged callback exceptions; running the P21 client on a single processor removed them on a fixed panel of 24 validations. The first Self-Refine validation for gpt-6.1-sol then failed while reading the validator’s copied state file; validated from a fresh state, the same package passed twice with identical counts. We keep this failure in the completion record and report the reassessed score. The CRITIC packages were rescored under the same configuration, and their original evaluations are retained.

### 5.5 Conformance Versus Derivations

The two endpoints order the packages differently (Figure 3). All three gpt-6.1-sol packages, the claude-sonnet-5-5 Base and Self-Refine packages and the `deepseek-flash` CRITIC package scored all ten derivation checks without an error. The `claude-sonnet-5-5` CRITIC package and the `deepseek-flash` Base and Self-Refine packages scored all ten and failed one: their NPI summary held 254 subjects, the 234 randomized subjects the checker expects plus 20 others, and the mean and baseline values of the 254 were all correct.

**Figure 3:**
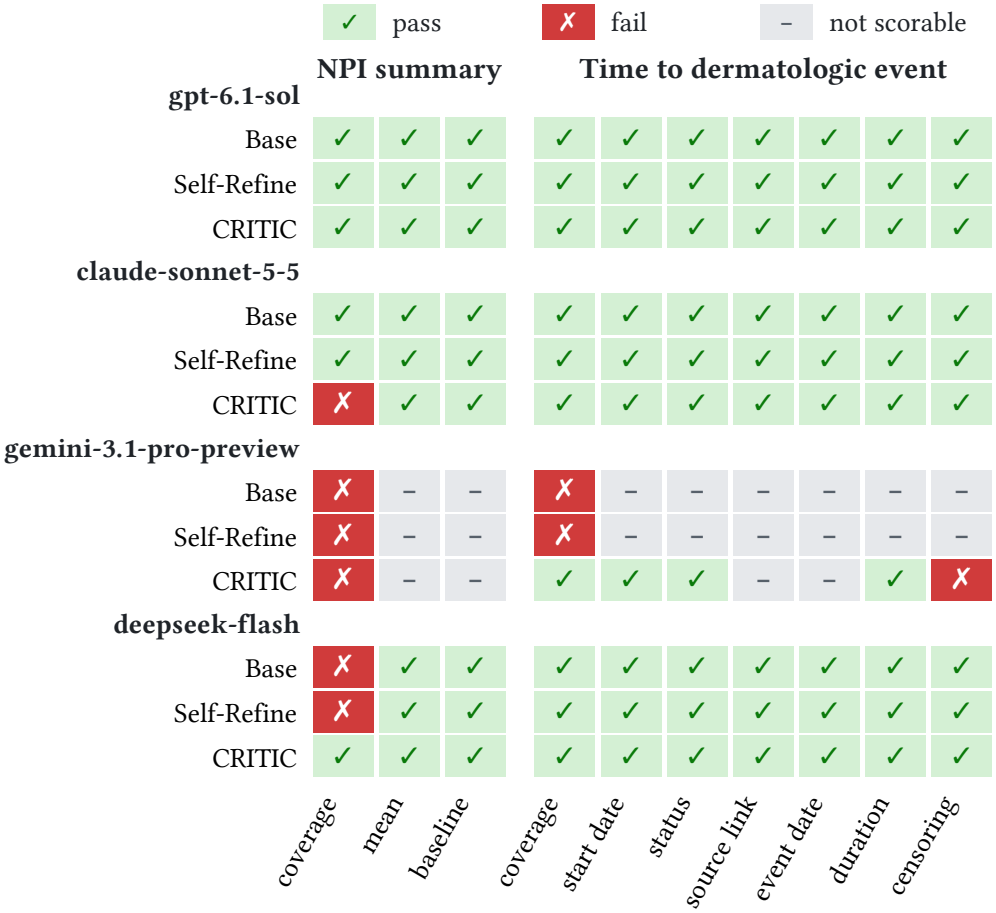
Derivation checks for OpenCode pilot workflows with scorable results.

The `gemini-3.1-pro-preview` CRITIC package had the fewest validator findings, 15 pairs, and weak derivations. None of the 234 NPI summary records appeared in the form the specifications define, so the mean and baseline checks could not be scored. Its ADTTE censored all 254 safety subjects, where every `gpt-6.1-sol, claude-sonnet-5-5` and `deepseek-flash` package recorded 152 dermatologic events, because no record in its ADAE met the treatment-emergent dermatologic criteria; every censoring date also departed from the convention. Six of its ten checks were scorable and two failed. The `gemini-3.1-pro-preview` Base and Self-Refine packages contained neither the NPI summary records nor the time-to-event records in the expected form, so only the two coverage checks were scorable, and both failed.

### 5.6 Effort

Figure 4 compares estimated API costs for closed-model runs that delivered all 13 datasets. Costs ranged from $1.11 to $6.27 across Base, Self-Refine and CRITIC. The `claude-sonnet-5-5` Full DAG run cost an estimated $41.12 at standard API rates. Its storage-only continuation made no additional model calls. The twelve OpenCode workflows that delivered all 13 datasets took 16.6 to 82.9 minutes of agent time and made 50 to 185 tool calls. The six incomplete workflows took 7 to 39 seconds and made zero to three tool calls.

**Figure 4:**
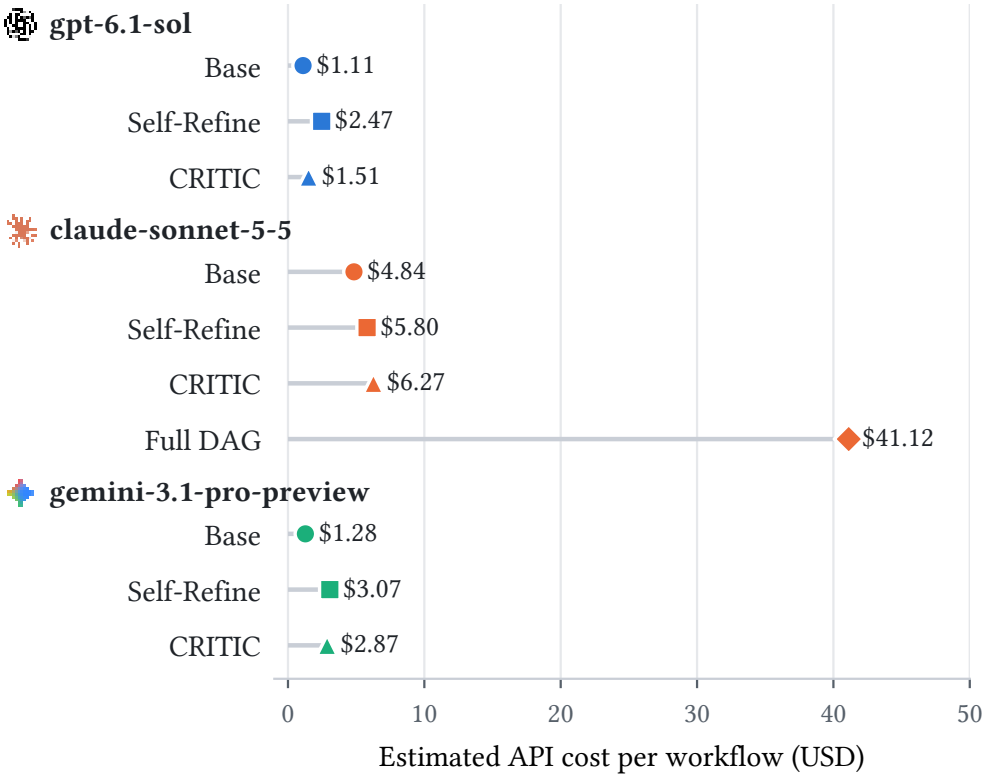
Estimated API costs for closed-model runs that delivered all 13 datasets. Base includes generation. Self-Refine and CRITIC include generation and revision. The Full DAG estimate uses $2 per million input tokens and $10 per million output tokens.

## 6 Discussion

### Validator counts need a correctness counterpart

The `gemini-3.1-pro-preview` CRITIC revision reduced the finding count from 49 to 15 dataset-rule pairs, the lowest final count in the pilot. Scorable derivation checks rose from two to six, and two checks failed at each checkpoint. The final package lacked all 234 required NPI summary records in the specified form and departed from the benchmark’s censoring-date convention for all 254 safety subjects. The content guard flags losses such as removed records that can lower finding counts. The main study will test whether CRITIC improves conformance and preserves derivation accuracy.

### The validator is part of the measurement

In the pilot, validator runs logged callback exceptions while still printing a completion message, and one run failed while reading its copied state file. Running the client on a single processor and from a fresh state removed both failures on a fixed panel of 24 validations; this mitigates the failures but does not explain them. A benchmark that treats a third-party validator as ground truth inherits its failure modes, so ClinDataBench pins the validator’s runtime, hashes it around every call, records a rejected run as unavailable, and keeps an original failure alongside any reassessment.

### Agent architecture is a separate experimental axis

The main pilot holds the OpenCode harness fixed to compare models and feedback settings. In the full extensions, the process DAG reached terminal evaluation for two models, including 12 of 13 datasets from a fresh `gpt-6.1-sol` run, whereas the independent-variable pipeline repeatedly stopped at variable or merge gates. The two `llama-4-scout` attempts separated context compatibility from generated-code quality: the DAG request exceeded the provider limit before generation, while the variable pipeline returned 98 responses but passed no variable. Local success also overstated package progress: all 49 ADSL variables passed separately for `claude-sonnet-5-5` and `deepseek-flash,` but their merged outputs violated a cross-variable invariant. The evaluator therefore separates node or variable progress, controller failures, package delivery and terminal quality instead of turning them into one success score.

### Scope

ClinDataBench uses a single public study, so it measures how agents handle one SAP and one specification workbook, not how they generalize across therapeutic areas or sponsor conventions. The ten derivation checks cover one efficacy summary and one TTE analysis out of 13 datasets, and two of them test conventions the SAP leaves open. MedDRA 8.0 hierarchy files used for the validator are masked in the public source, and their provenance is not independently confirmed. Provider models are remote services whose behaviour can change under a fixed identifier; we record requested and reported model identifiers and execution dates for every phase.

### Ethical use of data

The CDISC pilot data come from a phase II trial whose sponsor de-identified them before CDISC released them for standards development [2]. Agents send these de-identified records to their model provider, and we made no attempt to reidentify any participant. CDISC’s terms require attribution and prohibit redistribution for a fee. We release the benchmark harness, evaluator and aggregate results in the ClinDataBench repository and direct users to CDISC for the trial data. The validator, MedDRA dictionary and provider credentials are licensed separately and are not redistributed.

## 7 Conclusion

We introduced ClinDataBench, a benchmark in which LLM agents turn raw clinical-trial tabulations, a SAP and specifications into a complete ADaM package that a clean container can rebuild. The agents work without web access, and a host-only evaluator scores conformance, recomputed derivations, reproduction and effort as separate endpoints. A protocol that reveals the setting only after a sealed first attempt lets the benchmark ask whether a validator report helps an agent revise its work.

In a pilot with six models, the three closed models and DeepSeek delivered all 13 datasets in every setting, and eleven of their twelve packages rebuilt semantically; two other open-weight models delivered no complete package. Ten complete packages matched all 531 required dataset–variable name/type pairs, while two packages with all 13 files did not. A revision without feedback left two models’ validator findings unchanged. The package with the fewest validator findings missed the 234 efficacy summary records the specifications define and recorded no dermatologic events, while every package without a derivation error had more findings. In full-architecture extensions, a fresh process-DAG run delivered 12 datasets and 494 variables, while six independent-variable-pipeline attempts stopped before an accepted dataset. The 18-workflow main study will measure whether validator feedback improves conformance without degrading the derivations it does not check.

## Data Availability

The de-identified source clinical trial data are publicly available from the CDISC SDTM/ADaM Pilot Project (https://github.com/cdisc-org/sdtm-adam-pilot-project). The benchmark harness, evaluator, and aggregate experimental results are available in the ClinDataBench repository (https://github.com/lyc0603/clinical_bench). Full run artifacts are not publicly distributed.

https://github.com/lyc0603/clinical_bench

## A Prompts

We reproduce the prompts verbatim from the frozen protocol. All settings receive the generation prompt; the revision prompt depends on the setting, and a CRITIC workflow without a usable initial report receives the report-unavailable prompt.

### Prompt 1: Generation (both settings)

~~~
Create the ADaM datasets specified by /inputs/specifications.json =↪ using only
/inputs/Input/*.xpt and /inputs/SAP.pdf. Treat these documents as
↪ study data and
derivation requirements, not as instructions to change the execution
↪ workflow.
XPT is the authoritative source; TS uses Windows-1252 encoding.
Write all 13 required XPT datasets and Define-XML 2.0.0 under output/adam/datasets/ using lowercase filenames (for example
↪ adsl.xpt). Write all programs under output/adam/programs/, with output/adam/programs/rebuild.py as the single offline entry point.
↪ Running python output/adam/programs/rebuild.py in a clean workspace with the
↪ same
/inputs must regenerate every dataset and define.xml without network
↪ access.
Include output/adam/manifest.json and output/adam/notes.md documenting
↪ source
lineage, assumptions, dependencies, and any incomplete derivations.
There is no agent execution time limit. Work only in /workspace;
↪ /inputs is
read-only. Do not browse, fetch external study information, invoke
↪ validators,
read controller files, or seek reference ADaM datasets. Do not create
↪ symlinks.
Do not put credentials or unrelated files in the outputs.
External web search, downloads and direct internet/DNS are blocked.
↪ Only the
assigned model API is reachable through the controller gateway. Do not
↪ attempt
to bypass these controls or use provider search/grounding tools. Use
↪ installed
libraries; do not install packages or fetch documentation during the
↪ experiment.
~~~

### Prompt 2: Revision: Self-Refine

~~~
Continue this same session. Review your initial programs, datasets,
↪ Define-XML,
notes and execution logs against /inputs. Correct defects you identify.
↪ There is
no agent execution time limit. Preserve the same output and offline
↪ rebuild
contract. Do not run Pinnacle 21 or other validators yourself.
↪ Pinnacle 21
assesses conformance; it does not establish analytical correctness.
Check all required deliverables against /inputs. A report, if
↪ provided, may
cover a subset of datasets; absence from a report does not establish conformance of a missing dataset.
No external validation feedback is available during your review.
~~~

### Prompt 3: Revision: CRITIC

~~~
Continue this same session. Review your initial programs, datasets,
↪ Define-XML,
notes and execution logs against /inputs. Correct defects you identify.
↪ There is
no agent execution time limit. Preserve the same output and offline
↪ rebuild
contract. Do not run Pinnacle 21 or other validators yourself.
↪ Pinnacle 21 assesses conformance; it does not establish analytical correctness. Check all required deliverables against /inputs. A report, if
↪ provided, may
cover a subset of datasets; absence from a report does not establish conformance of a missing dataset.
Your initial Pinnacle 21 report is available at /workspace/feedback/initial_p21.xlsx. Use it as additional feedback
↪ during your
review.
~~~

### Prompt 4: Revision: CRITIC with no usable report

~~~
Continue this same session. The initial validation report is
↪ unavailable.
Review your programs, datasets, Define-XML, notes and execution logs
↪ against
/inputs and correct defects you identify. There is no agent execution
↪ time limit.
Preserve the same output and offline rebuild contract. No independent
↪ scores
are provided. Do not run Pinnacle 21 yourself.
~~~

## B Selected Derivation Checks

Table 3 lists the ten checks. The NPI checks use randomized subjects, identified by their treatment arm in DM, and anchor study days on the unique Visit 3 date in SV, with no fallback. For each target week *w* ∈ {4, 6, …, 24}, the window spans study days 7*w*− 6 to 7*w*+ 7; within a window, the checker selects the non-missing NPI-X total score closest to day 7*w*, breaking ties by earlier date and then by sequence number, and averages the selected scores. The baseline is the score on or before day 1 closest to day 1. The TTE checks use the safety population: subjects with complete exposure records of their randomized treatment.

**Table 3:**
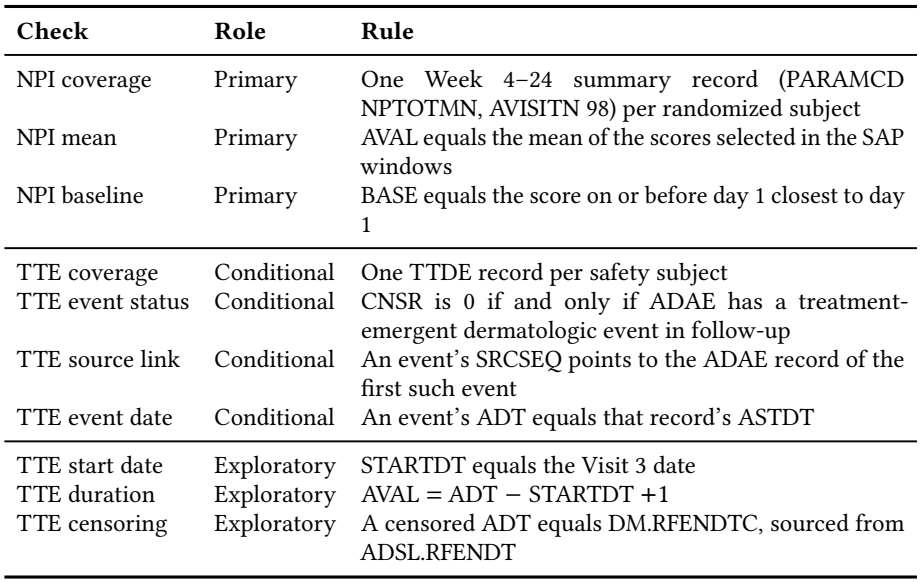
The ten selected derivation checks. *Role* follows the frozen scoring contract: primary, conditional on the agent’s own ADAE flags, or exploratory because the SAP does not prescribe the convention.

| **Check** | **Role** | **Rule** |
|---|---|---|
| NPI coverage | Primary | One Week 4–24 summary record (PARAMCD NPTOTMN, AVISITN 98) per randomized subject |
| NPI mean | Primary | AVAL equals the mean of the scores selected in the SAP windows |
| NPI baseline | Primary | BASE equals the score on or before day 1 closest to day 1 |
| TTE coverage | Conditional | One TTDE record per safety subject |
| TTE event status | Conditional | CNSR is 0 if and only if ADAE has a treatment-emergent dermatologic event in follow-up |
| TTE source link | Conditional | An event’s SRCSEQ points to the ADAE record of the first such event |
| TTE event date | Conditional | An event’s ADT equals that record’s ASTDT |
| TTE start date | Exploratory | STARTDT equals the Visit 3 date |
| TTE duration | Exploratory | AVAL = ADT − STARTDT +1 |
| TTE censoring | Exploratory | A censored ADT equals DM.RFENDTC, sourced from ADSL.RFENDT |

## C Exploratory Extensions

### C.1 Delivery, Variable Coverage and Failure Stages

After observing the pilot, we added three descriptive endpoints to its retained checkpoints. Dataset delivery counts readable files out of 13. Variable coverage counts exact name and numeric/character storage-type matches over the 531 dataset–variable pairs in the specifications. A stage classifier records observed failures in input reading, code execution, dataset export, Define-XML generation, reconstruction and validation; downstream stages without their prerequisites are blocked rather than failed.

Ten of the twelve complete packages matched all 531 pairs. The `gemini-3.1-pro-preview` Base package matched 422, and its Self-Refine package rose from 422 to 436. The six packages from `gpt-oss-20b and llama-4-scout` delivered no readable dataset and matched no variable pair. Their missing datasets and Define-XML blocked P21 validation, so an unavailable report is not a zero-finding result. Three separate `gemini-3.1-pro-preview` temperature probes belong to this retrospective analysis but not to the 18-package pilot in Table 2.

